# Nonlinear Classification of EEG recordings from patients with Alzheimer’s Disease using Gaussian Process Latent Variable Model

**DOI:** 10.1101/2020.05.07.20093922

**Authors:** S. Rajintha. A. S. Gunawardena, Fei He, Ptolemaios Sarrigiannis, Daniel J. Blackburn

**Affiliations:** Centre for Data Science, Coventry University, Coventry CV1 2JH, UK (,); Academic Unit of Radiology at the University of Sheffield, Sheffield S1 2TN, UK; Department of Neuroscience, University of Sheffield, Sheffield S10 2HQ, UK

## Abstract

In this work, nonlinear temporal features from multi-channel EEGs are used for the classification of Alzheimer’s disease patients from healthy individuals. This was achieved by temporal manifold learning using Gaussian Process Latent Variable Models (GPLVM) as a nonlinear dimensionality reduction technique. Classification of the extracted features was undertaken using a nonlinear Support Vector Machine. Comparisons were made against the linear counterpart, Principle Component Analysis while exploring the effect of the time window or EEG epoch length used. It was demonstrated that temporal manifold learning using GPLVM is better in extracting features that attain high separability and prediction accuracy. This work aims to set the significance of using GPLVM temporal manifold learning for EEG feature extraction in the classification of Alzheimer’s disease.

## I. Introduction

Dementia due to Alzheimer’s disease (AD) is a neurodegenerative illness that mainly occurs in elderly individuals and evolves over a prolonged period of time. The diagnosis of neurological disorders such as AD in early stages and the accurate disease progress characterisation can be vital for the treatment and improvement of patient’s life [1, 2]. However, the current diagnostic methods depend mainly on clinical history, mental status examinations and neuroimaging scans [2]. These methods are expensive, time-consuming and sometimes inaccurate [3]. Thus, a cost-effective precise diagnosis method is needed, especially for the early detection of AD, the most common neurodegenerative disorder [4]. One such method that is non-invasive and economical, has no-contradictions and previously widely researched is the electroencephalogram (EEG) [5].

The EEG, even though recorded at the scalp level, reflects the grossly summed currents of the electrical fields generated by the neural activity in the cortical neural circuits. Therefore, through the EEG, the behaviour and integrity of underlying neural circuits can thus be indirectly studied [8]. Slowing of EEG rhythms, loss of signal complexity and changes in the strength of synchronisation between pairs of recording electrodes are some of the abnormalities previously observed in AD patients [12, 13].

Previous work in dementia patients [6, 7] investigated the nonlinear dynamics of EEG measures, and showed that the computed global brain electrical nonlinear indices exhibited specific patters of dysfunction. In the case of AD, when analysing the EEG nonlinear dynamics, researchers have demonstrated a reduction in nonlinear complexity [9, 10]. A comprehensive review of the nonlinear dynamical analysis of EEG measures [11] was recently produced and sheds light on the importance of this type of approach for the diagnosis of neurodegenerative diseases.

Numerous EEG studies have previously revealed the importance of nonlinear methods for the diagnosis of AD. These range from time-series methods, spectral analysis and machine learning techniques [11, 13 - 15].

EEG recordings are high-dimensional data (i.e. number of channels and high-frequency sampling time points) and as such directly applying standard classification methods to the raw EEG data can be problematic. Therefore, how to select the important nonlinear features in a reduced lower-dimensional space is an important but also challenging problem. The corresponding comparison with classical linear dimension reduction methods is also lacking.

In this work, we explore the unsupervised Gaussian Process Latent Variable Model (GPLVM) [19], which models the joint distribution of the observed data and their corresponding representation in a low dimensional latent space. We compared the linear and nonlinear dimensionality reduction approaches, i.e. Principle Component Analysis (PCA) and GPLVM, for the classification of EEG recordings of patients with AD, and demonstrated the advantages from utilising a nonlinear approach.

This paper is organised as follows section II and III briefly introduce Gaussian Process modelling and GPLVM respectively. Section IV provides specifics of the EEG data used, pre-processing steps and how GPLVM was used for classification. Section V discusses the results obtained followed by the concluding remarks while section VI discusses the future work.

## II. Gaussian Process

Gaussian Process (GP) has been widely used as a Bayesian non-parametric model in various machine learning applications [16]. A GP is a distribution over functions where a finite set of random function variables, ***f*** = [*f*(***x***_1_),…, *f*(***x****_N_*)]^T^ where *x_i_* ϵ ℝ^1×^*^D^* is the *i*^th^ sample of a *D* dimensional input, are modelled as a joint multivariate Gaussian distribution with mean ***µ*(*X*)** and covariance matrix ***K*(*X, X*)**, in which the data matrix ***X*** = [***x***_1_,…, ***x****_N_*]^T^ and

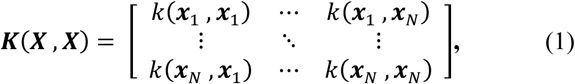

is a positive semidefinite matrix, where *k*(***x****_i_*,***x****_j_*) is the kernel function, also called the covariance function with hyperparameters ***θ***. ***µ*(*X*)** = [*m*(***x***_1_), …,*m*(***x****_N_*)]^T^, where *m*(***x****_i_*) is the mean function. It is common to have the mean function *m*(***x****_i_*) = 0. Therefore, assigning a multivariate Gaussian prior over *f*, 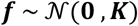.

Given ***y****_i_* = *f*(***x****_i_*) + *ε_i_*, where *ε* ~ unfigure 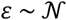 (*0,σ*^2^) and ***Y*** = [ ***y***_1_, …, ***y****_N_*]^T^. Considering a Gaussian likelihood where 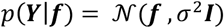, ***I*** ϵ ℝ*^N^*^×^*^N^* is an identity matrix. Therefore, given a set of new input samples ***X***_*_ = [***x***_*1_,…, ***x***_*_*_N_*,]^T^, the inference for new outputs or observations ***Y***_*_ = [***y***_*1_,…, ***y***_*_*_N_*,]^T^ are given by multivariate Gaussian distribution, thus 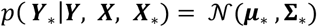 where

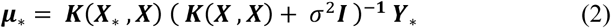

is the mean of the distribution of the new observations and

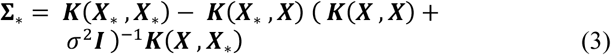

defines its covariance matrix. It should be noted that if the covariance function used is one which allows non-linear functional mappings between ***X*** and ***Y***, e.g. the Radial Basis Function (RBF) kernel, then the GP model provides a probabilistic non-parametric nonlinear model and vice-versa for a linear covariance function.

The hyper-parameters ***θ*** of the covariance function *k*(***x****_i_*, ***x****_j_*) can be extended to include the noise variance *σ*^2^. The hyper-parameters are then determined by the maximisation of the marginal log-likelihood

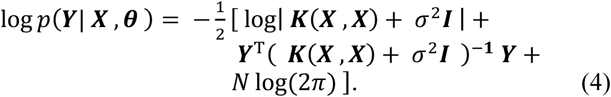

The optimisation of the hyper-parameters are guided by the gradients of the marginal log-likelihood with respect to each hyper-parameter. For an in depth understanding of GPs refer to [17, 18].

## III. Gaussian Process Latent Variable Model

A GPLVM [19] assumes that the observed dataset ***Y*** is generated from a lower dimensional data ***X***. Therefore, the GPLVM framework learns the mapping of a high-dimensional dataset ***Y*** ϵ ℝ*^N^* ^×^ *^D^* and the corresponding low dimensional latent space ***X*** ϵ ℝ*^N^* ^×^ *^D^*, *Q < D*, using a GP mapping from the latent space to the data space, ***X*** → ***Y***. Let ***Y*** = [***y***_1_,…, ***y****_N_*]^T^, *y_i_* ϵ ℝ^1 ×^ *^D^*, and ***X***= [*x*_1_,…,***x****_N_*]*^T^, x_i_* ϵ ℝ^1 ×^ *^Q^*. Given a covariance function for the GP, *k*(***x****_i_*, ***x****_j_*), the likelihood *p*(***Y|X,θ***) of the data given the latent positions is

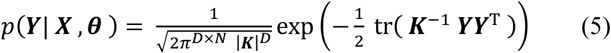

where ***K* = *K*(*X, X*)** as defined in Equation (1) and *θ* is the respective hyper-parameters of the covariance function which also includes the noise variance of the data. Thus the maximising the marginal log-likelihood, log *p*(*Y|X*, *θ*), with respect to ***X*** and ***θ***, the optimal estimates 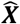 and 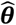 are obtained.

In [20] the authors provide a probabilistic formulation for PCA. In GPLVM the use of a linear covariance function, restricting the mapping ***X*** → ***Y*** to linearity, consequently results in an alternative probabilistic model for PCA as given by [20]. However, the use of a covariance function that allows for nonlinear functional mappings provides a probabilistic nonlinear latent variable model. Therefore GPLVM is a nonlinear manifold learning technique for nonlinear dimensionality reduction.

### A. Dissimilarity Preservation

The GPLVM ensures a smooth mapping from the latent to the data space. However, the mapping does not guarantee that the local structures within the data space will be preserved in the latent space. Conversely, this mapping ensures that two points which are ‘distant’ in the data space cannot be placed closer together in the latent space. This implies a discontinuity in the mapping. Thus, in a certain sense the GPLVM is dissimilarity preserving [33].

## IV. Materials and Methods

All participants were prospectively selected in this study and provided informed consent while this project was approved by the Yorkshire and the Humber (Leeds West) Research Ethics Committee (reference number 14/YH/1070). Patients were recruited in the Sheffield Teaching Hospital memory clinic, a young-onset memory clinic that deals with people mostly aged under 65. All AD EEG data and most Healthy Controls (HC) have been previously used in a recent study by Blackburn *et al*. in [21]. Task-free EEG recordings that requires minimal cooperation of AD patients were used; typically this group of patients can have difficulty engaging with and following cognitive tasks. The HC participants were enrolled through opportunistic sampling and word of mouth. The following sub-sections A. and B. summaries the details of the study design used in [21] to acquire the EEG recording of AD and HC groups.

### A. Patients and Healthy Controls

Diagnosed between 1 month to 2 years prior to the EEG recordings, participants with AD were in the mild to moderate stages of the disease, with a mean Mini Mental state Examination (MMSE) score of 20.1 (standard deviation 4.00). The diagnosis was reached by taking into account clinical history, neurological examination, neuropsychological scores, and neuro-radiological findings. The determination of the cognitive profile of AD patients and HC was achieved using a series of extensive neuropsychological tests, appropriately tailored to be sensitive to cognitive difficulties typically seen in AD. To exclude other alternative causes of dementia all patients were scanned with high-resolution structural MRI.

The HC were of age and gender matched and everyone underwent a series of extensive neuropsychology testing, and structural and functional MRI imaging that were normal.

EEG recordings from a total of 16 AD patients and 12 HC participants were used in this study. The specific details of the neuropsychology tests and the functional MRI scans of the AD cohort are described in great detail in previous work [21].

### B. EEG Data Acquisition

All EEG data was acquired with an XLTEK 128-channel headbox (Optima Medical LTD, Surrey, UK) at a sampling rate of 2000 Hz with scalp Ag/AgCL electrodes. A modified 10/10 overlapping a 10/20 international system of electrode placement method was utilised to acquire the EEG recordings.

The recordings were undertaking with a referential montage were a linked earlobe reference was used (jump cables were devised to combine the right and left earlobe electrodes while impedances where kept equal between sides). Subsequently the following 23 bipolar channel pairs were produced for the analysis; F8-F4, F7-F3, F4-C4, F3- C3, F4-FZ, FZ-CZ, F3-FZ, T4-C4, T3-C3, C4-CZ, C3-CZ, CZ-PZ, C4-P4, C3-P3, T4-T6, T3-T5, P4-PZ, P3-PZ, T6- O2, T5-O1, P4-O2, P3-O1 and O2-O1.

During the EEG recording the participants were encouraged not to think about anything specific. With alternating 5 minute Eye-Open (EO) and Eye-Close (EC) epochs, 30 minute resting state EEG recordings were obtained from all participants while they were prompted if demonstrated any signs of drowsiness. All EEGs were reviewed by an experienced neurophysiologist using the XLTEK review station with time-locked video recordings (Optima Medical LTD). From the 30-minute resting state EEG recordings, 12 second artefact free EO and EC epochs were isolated. Subsequently, for each of the EO and EC epochs, mini-epochs of 2, 3 and 4 seconds were produced.

### C. Pre-processing tasks

In the analysis of EEG signals, the use of digital filtering is of common practise. Usually in EEG signal processing the application of a high-pass filter to attenuate low frequency artefacts relating to eye blinking and slow movements is used. Furthermore, low-pass filtering is used to filter out higher frequencies and a notch filter is often applied to filter out the frequency component relating to the AC power line voltage [22].

Even though filters are extremely useful in attenuating certain frequency components from the signal of interest, filtering is not entirely harmless. Several studies have demonstrated that filtering can introduce distortions in the temporal structure of EEG signals [23-26]. This would not be much of an issue if only the magnitude of the frequency spectrum of the signal is used. However, in this study, since the high-dimensional temporal structure of the multichannel EEG is examined, the use of filters would pose a major issue due to the distortions induced. Thus, the Fast Fourier Transform (FFT) was implemented to remove unwanted frequency components. Thereafter, the inverse-FFT was used to reconstruct the time domain signal without the unwanted frequency components.

The following frequency components were removed; 0 - 2Hz which could relate to eye-blinks and slow movement artefacts and all frequencies above 100Hz that could be EMG related activity. Thus the analysis in this work was conducted using EEG frequencies between 2 - 100Hz. After removing the unwanted frequency components the time-domain signal was recovered via the inverse FFT. This was then down sampled to 200Hz.

### D. Low-Dimensional Manifold Learning of High-Dimensional Multi-channel EEG Using GPLVM

As mentioned in Section IV.B, the number of EEG channels for each participant included in the analysis was 23. The low-dimensional latent space for each AD and HC subject is thus obtained by reducing the temporal dimension via nonlinear dimensionality reduction using the GPLVM framework mentioned in section III. Therefore, from the definition of the data space ***Y*** ϵ ℝ*^N^* ^×^ *^D^* in section III, *N* = 23 and *D* is the temporal dimension that is to be reduced. Once dimensionality reduction is done, the respective latent space ***X*** for each AD and HC subject will be such that *X* ϵ ℝ*^N^* ^×^ *^Q^*, where the latent dimension *Q* is set as *Q =* 2. The GPLVM is initialised using PCA. The heavy tailed Matérn 12 covariance function was chosen over the commonly used RBF kernel. This is because nearly a 100% recover accuracy was obtained in the GPLVM mapping ***X*** → ***Y*** when using the Matérn 12 covariance function. The initialisation of the hyper-parameters of the covariance function was done in a trial and error fashion in order to obtain favourable classification accuracies.

Before applying dimensionality reduction each and every EEG channel of all subjects were de-meaned and normalised such that the absolute maximum value attained by the respective channels is 1.

### E. Classification

In order to demonstrate the classification ability using the low-dimensional latent spaces of each EEG channel, binary classification using non-linear Support Vector Machine [27] with a RBF kernel was used. The two latent dimensions (*Q* = 2 as mentioned earlier) corresponding to each channel were used as the features of that respective channel. Thus, 23 separate binary classifications using the respective low-dimensional latent spaces as features of the 23 EEG channels were carried out. The non-linear SVM classification was implemented in Python using the Scikit-learn package.

The respective latent spaces of the high-dimensional EEG data were produced for all AD and HC subjects using GPLVM. Out of the 16 AD patients and 12 HC subjects, the respective latent spaces of 10 AD and 10 HC subjects were used to train the 23 nonlinear SVM classifiers. The remaining latent spaces of 6 AD and 2 HC subjects were used to test the prediction accuracies of the respective classifiers.

## V. Results

In Fig. 1 - 8 illustrated in this section, the triangular markers relate to the respective testing set of features and the circular markers relate to the training set. All orange markers indicate features of AD and light blue indicates HC. The orange shaded regions relate to the area of the EEG channel latent spaces in which AD features are to be expected and similarly the blue shaded regions for HC.

**Figure 1.**
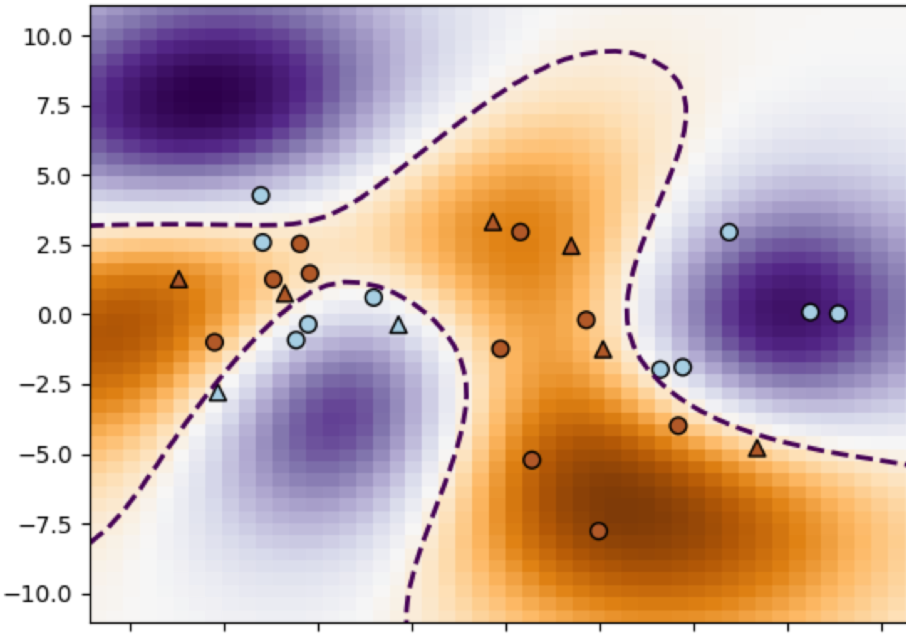
EC GPLVM Latent space of channel P3 – O1, that attained the best prediction accuracy 100% within the 3 second mini-epoch that immediately follows eye closure (0-3 time window). SVM score during training 0.95. Orange markers indicate features of AD and light blue indicates HC. The triangular markers denote the testing set of features and the circular markers denote training set.

**Figure 2.**
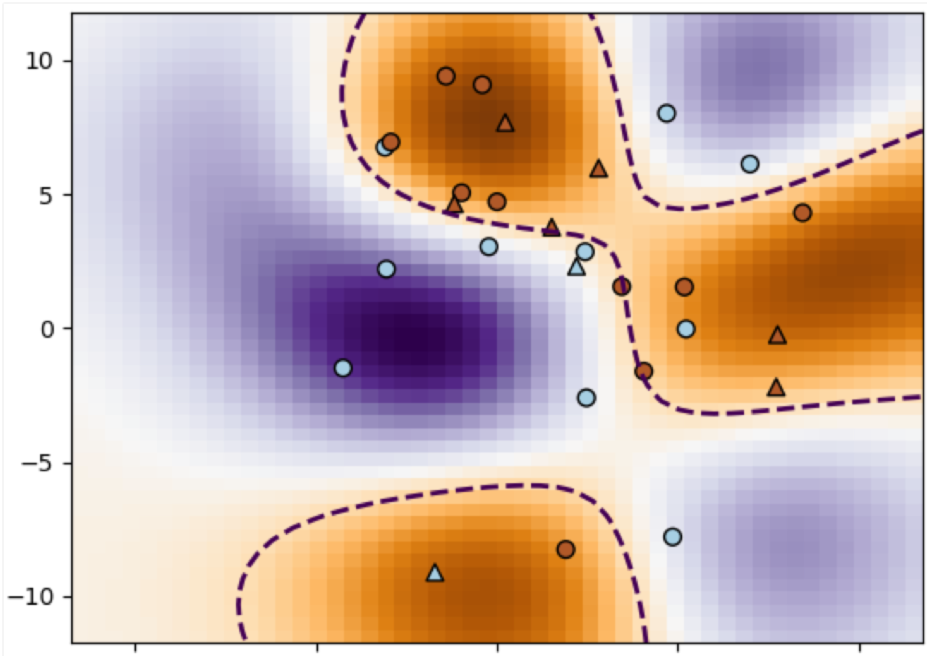
EC GPLVM Latent space of channel P4 - PZ, that attained the 2nd best prediction accuracy of 87.5% within the 0-3 time window. SVM score during training 0.85

**Figure 3.**
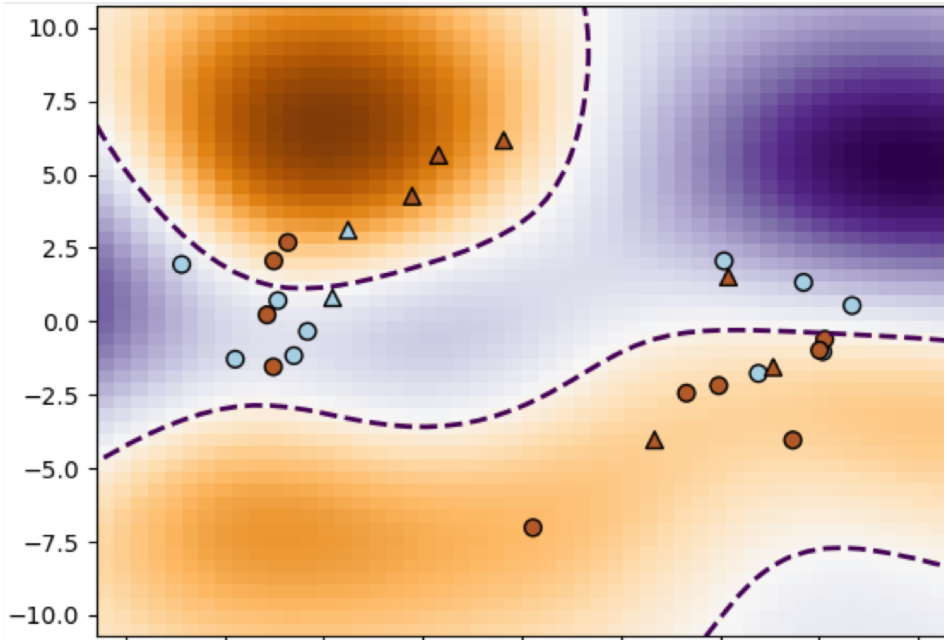
EC PC A Latent space of channel CZ - PZ, that attained the best prediction accuracy 75% within the 0-3 time window. SVM score during training 0.8

**Figure 4.**
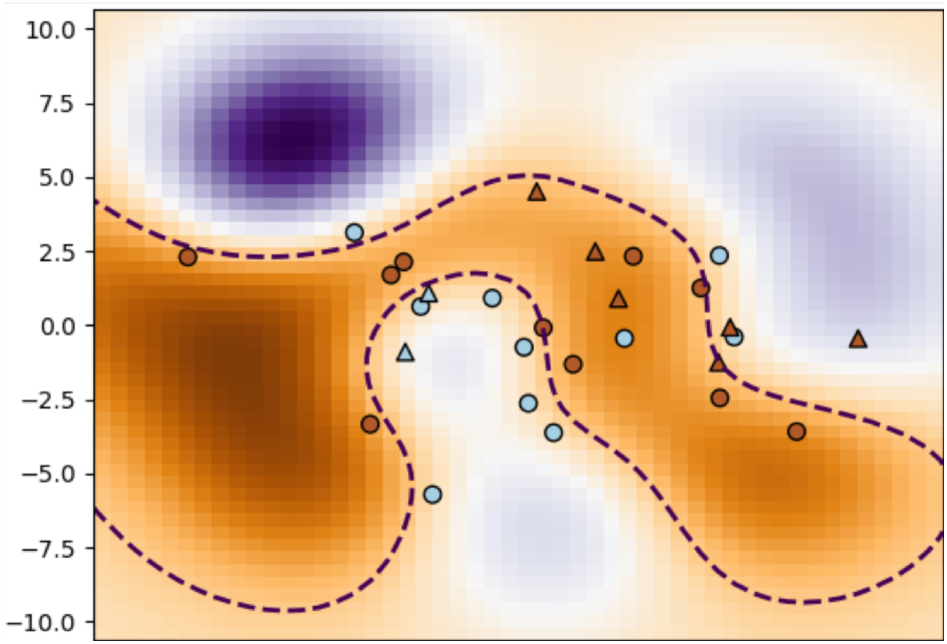
EC PCA Latent space of channel F3 - C3, that attained the 2^nd^ best prediction accuracy of 75% within the 0-3 time window. SVM score during training 0.95

**Figure 5.**
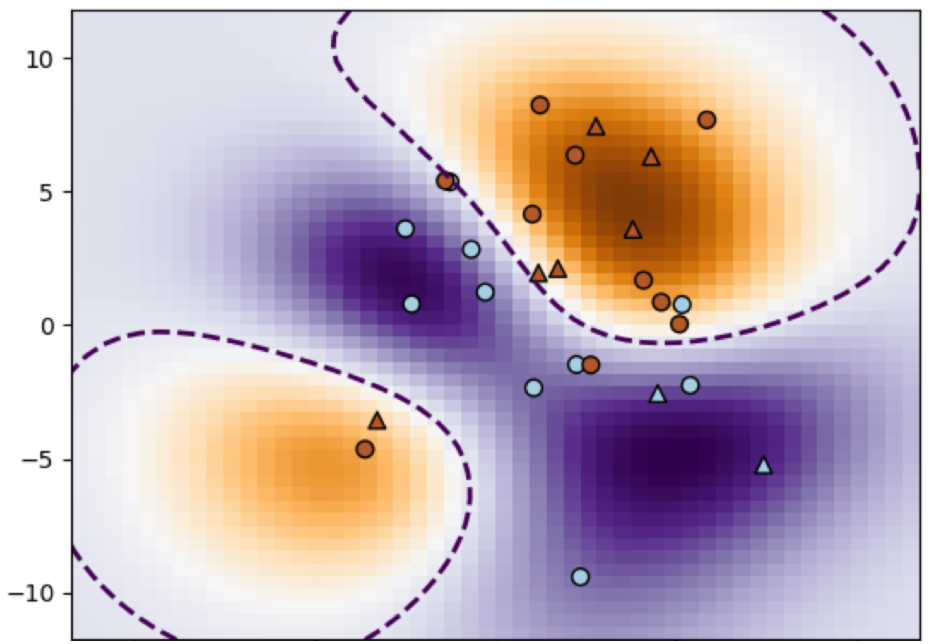
EO GPLVM Latent space of channel T6 - 02, that attained the best prediction accuracy 100% within the 0-3 time window. SVM score during training 0.85

**Figure 6.**
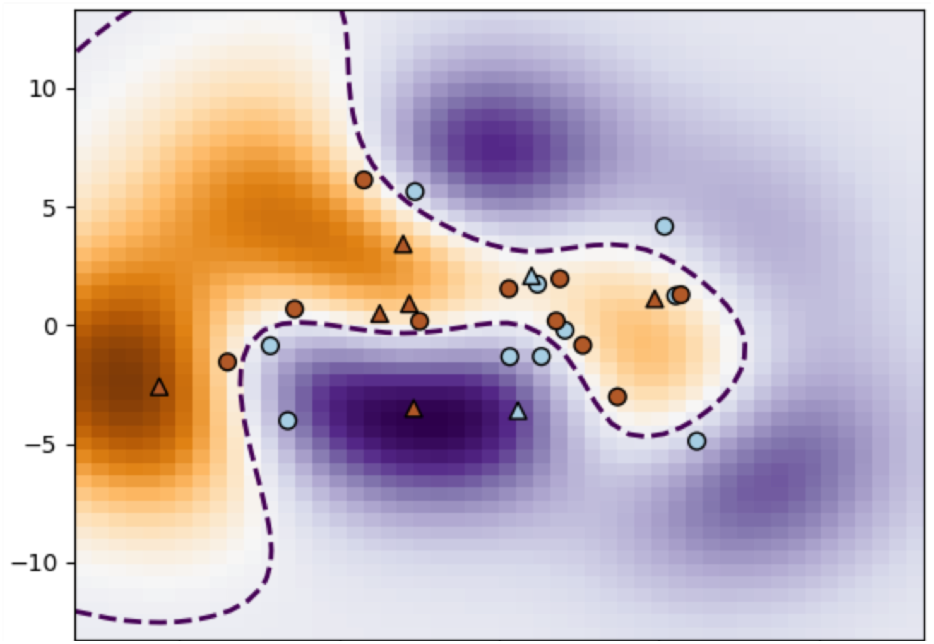
EO GPLVM Latent space of channel F4 - C4, that attained the 2nd best prediction accuracy of 75% within the 0-3 time window. SVM score during training 0.85

**Figure 7.**
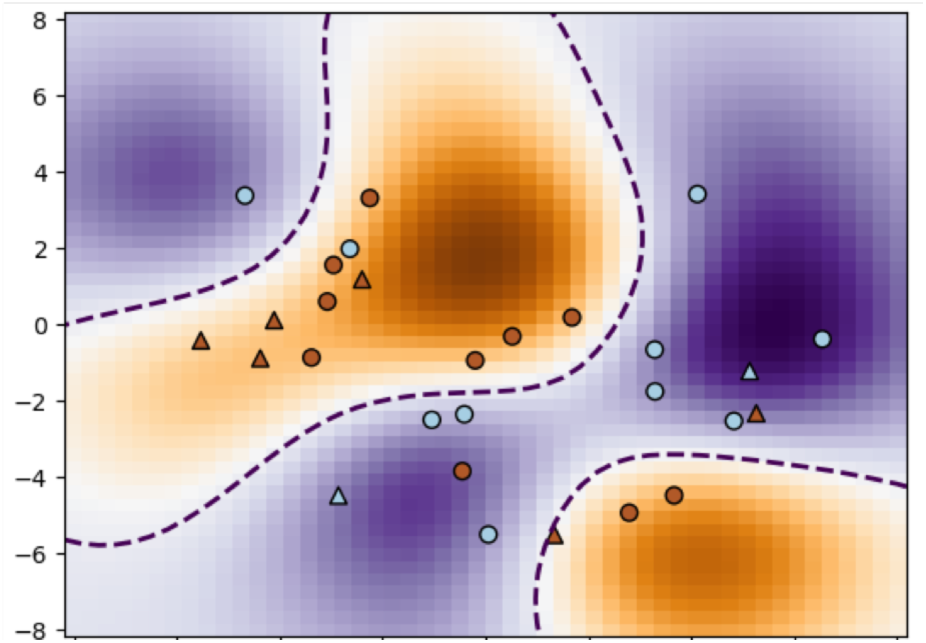
EO PCA Latent space of channel P3 – O1, that attained the best prediction accuracy 87.5% within the 0 – 3 time window. SVM score during training 0.9

**Figure 8.**
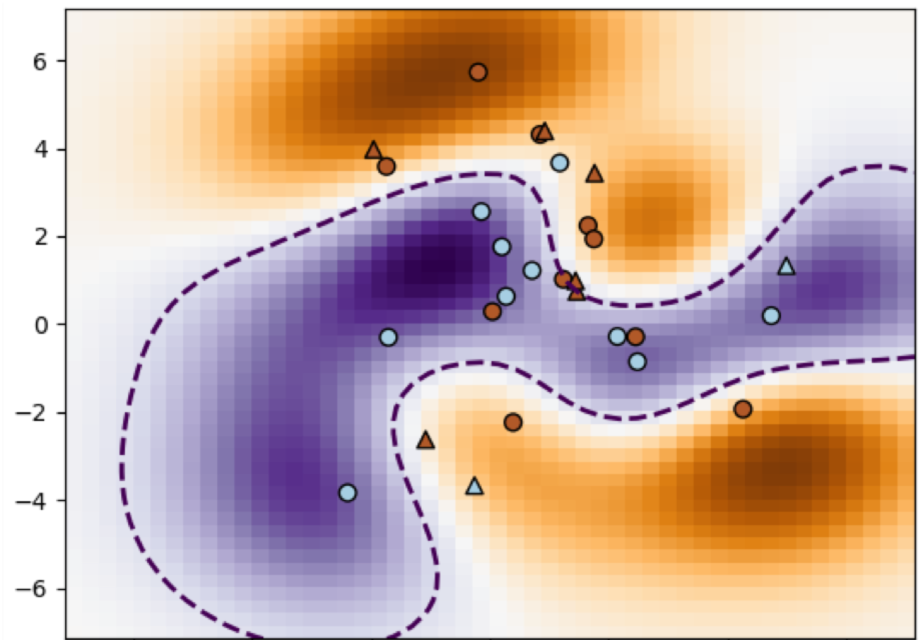
EO PCA Latent space of channel F4 – FZ, that attained the 2nd best prediction accuracy of 87.5% within the 0 – 3 time window. SVM score during training 0.8

The complete process of feature extraction using dimensionality reduction and classification was done for three different lengths of time windows (epoch lengths). This was done in order to see how GPLVM performs against PCA for different epoch lengths. Furthermore, in the results below, ‘prediction accuracy’ means how well the trained classifier predicts the classification (AD or HC) of the testing features. This does not account for how well the features used for training is classified. The SVM score indicates how well the training features can be separated in the latent space of the corresponding EEG channel.

Tables 1 and 3 below summarises the prediction accuracy results for all the 23 classifiers relating to the 23 EEG channels, when PCA is used for dimensionality reduction, in EC and EO EEG resting states respectively. The tables show the mean prediction accuracy across all the channels. Also shown is the best prediction accuracy attained and the corresponding channel which attains it out of all the other channels. Similarly, Tables 2 and 4 is when GPLVM is used for dimensionality reduction in the EC and EO EEG resting states respectively. Tables 2 and 4 also show the corresponding hyper-parameter initialisations used for the covariance function.

**Table 1.** FEATURES EXTRACTED USING PCA - EC

| PCA Classification Accuracy % |  |  | Time Window |
| --- | --- | --- | --- |
| Mean | Best | Best channel |  |
| 53.260 | 100 | P4 – O2 | 0 – 2 |
| **51.63** | **75** | **CZ – PZ** | **0 – 3** |
| 54.89 | 87.5 | T6 – O2 | 0 – 4 |

**Table 2.** FEATURES EXTRACTED USING GPLVM - EC

| GPLVM Classification Accuracy % |  |  | Covariance Function Hyper-parameters initialisations |  | Time Window |
| --- | --- | --- | --- | --- | --- |
| Mean | Best | Best channel | Variance | Length-scale |  |
| 59.78 | 100 | T6 – O2 | 0.5 | 0.5 | 0 – 2 |
| **50** | **100** | **P3 – O1** | **1** | **1** | **0 – 3** |
| 59.78 | 100 | T6 – O2 | 0.5 | 1 | 0 – 4 |

**Table 3.** FEATURES EXTRACTED USING PCA - EO

| PCA Classification Accuracy |  |  | Time Window |
| --- | --- | --- | --- |
| Mean | Best | Best channel |  |
| 48.91 | 87.5 | F4 – FZ | 0 – 2 |
| **53.80** | **87.5** | **P3 – O1** | **0 – 3** |
| 52.17 | 75 | C4 – CZ | 0 – 4 |

**Table 4.** FEATURES EXTRACTED USING GPLVM - EO

| GPLVM Classification Accuracy |  |  | Covariance Function Hyper-parameters initialisations |  | Time Window |
| --- | --- | --- | --- | --- | --- |
| Mean | Best | Best channel | Variance | Length-scale |  |
| 60.87 | 100 | T3 – T5 | 2 | 0.5 | 0 – 2 |
| **55.43** | **100** | **T6 – O2** | **1.5** | **1** | **0 – 3** |
| 59.24 | 87.5 | CZ – PZ | 1 | 10 | 0 – 4 |

It is evident from the results shown in the following subsections that when GPLVM is used as a dimensionality reduction technique for feature extraction, in both EC and EO resting state EEGs, across all time windows, at least one channel attains a prediction accuracy of 100%. This is true with the exception of one case when the time window 0 - 4 is used in the EO instance. Furthermore, out of all the time widows used, the 0-3 time window is of a special case. This is because in this time window, while a 100% prediction accuracy is attained at least by one channel, this particular time window also attains high SVM scores for the respective best channel for both EO and EC instances. This indicates that this time window may be of particular use when further analysing the EEG. This is particularly true in the EC case, as shown in Fig. 1 and Fig. 2.

### A. Eye-Close results

### B. Eye-Open results

## VI. Conclusion and Future Works

In this work, purely temporal based features are used for the classification of AD and HC resting state EEGs in both EC and EO instances. The high-dimensional multichannel EEG is projected to a nonlinear low-dimensional manifold and a nonlinear mapping from the data space to the latent space by means of GPLVM. A comparison was done using PCA which is the linear counterpart to GPLVM. From the results it is evident that nonlinear dimensionality reduction using GPLVM provides comparably better results. Also, certain interesting findings about the time window used for the analysis was made in regard to both the prediction accuracy of new features as well as the feature separability in the latent space of EEG channels.

Even though certain interesting findings were made there is much room for further investigation into the use of GPLVM for temporal feature extraction in EEGs. GPLVM does not preserve the local structures present in the data in the latent space and only preserves global dissimilarities. Thus, some important local features may be lost. Using back constraint GPLVM proposed in [33], a compromise can be made between local similarity preservation and global dissimilarity preservation. Furthermore, GPLVM is a static model, thus the temporal EEG manifold is regarded as static. Incorporating the temporal dynamics using GP Dynamic Models [34] would prove to be useful. Also, one important aspect that needs to be added to this work is a neurological interpretation of the result. Especially in regard to the channels that attain 100% prediction accuracies and high SVM scores in both EC and EO cases as well as the 0-3 time window.

One important future work is to further investigate more detailed form of the nonlinearity using nonlinear dynamic modelling [28, 29] and causality measures in both time and frequency domain [30-32],

## Data Availability

Currently, we have no plan to make the data publicly available. However, some data may be available if consent/agreement has been made with our clinical co-authors and corresponding registration procedure has been done.

## Acknowledgment

SRASG and FH would like to acknowledge Coventry University for providing the Trailblazer PhD studentship. The EEG data was funded by a grant from the Alzheimer’s Research UK, grant reference number ARUK-PPG20114B-25. We would like to thank all the patients and volunteers who took part in this study. This is a summary of independent research carried out at the NIHR Sheffield Biomedical Research Centre (Translational Neuroscience). The views expressed are those of the author(s) and not necessarily those of the NHS, the NIHR or the Department of Health.

